# Manual Chest PRESSURE during Direct Current Cardioversion for Atrial Fibrillation: A Randomised Control Trial (PRESSURE-AF)

**DOI:** 10.1101/2023.12.05.23299530

**Authors:** David Ferreira, Philopatir Mikhail, Joyce Lim, Max Ray, Jovita Dwivedi, Stephen Brienesse, Lloyd Butel-Simoes, William Meere, Adam Bland, Niklas Howden, Michael Malaty, Mercy Kunda, Amy Kelty, Michael McGee, Andrew Boyle, Aaron L Sverdlov, Maged William, John Attia, Nicholas Jackson, Gwilym Morris, Malcolm Barlow, James Leitch, Nicholas Collins, Thomas Ford, Bradley Wilsmore

## Abstract

**Background:** Direct current cardioversion is frequently used to return patients with atrial fibrillation (AF) to sinus rhythm. Chest pressure during cardioversion may improve the efficacy of cardioversion through decreasing transthoracic impedance and increasing cardiac energy delivery. We aimed to assess the efficacy and safety of upfront chest pressure during direct current cardioversion for atrial fibrillation with anterior-posterior pad positioning.

**Design, Setting and Participants:** This was a multi-center, investigator-initiated, patient and analysis blinded, randomised clinical trial. Recruitment occurred from 2021 to 2023. Follow-up was until hospital discharge. Recruitment occurred across three centers in New South Wales, Australia. Inclusion criteria were age ≥18, referred for cardioversion for AF, and anticoagulation for three weeks or transoesophageal echocardiography excluding left atrial appendage thrombus. Exclusion criteria were other arrhythmias requiring cardioversion, such as atrial flutter and atrial tachycardia.

**Intervention and Outcomes:** The intervention arm received chest pressure during cardioversion from the first shock. The primary efficacy outcome was total joules required per patient encounter. Secondary efficacy outcomes included first shock success, transthoracic impedance, cardioversion success and sinus rhythm at 30 minutes post cardioversion.

**Results:** 311 patients were randomised, 153 to control and 158 to intervention. There was no difference in total joules applied per encounter in the control arm versus intervention arm (356.4 ± 301 vs 413.8 ± 347, P=0.25). There was no difference in first shock success, total shocks provided, average impedance and cardioversion success.

**Conclusions and Relevance:** This study does not support the routine application of chest pressure for direct current cardioversion in atrial fibrillation. Reducing the complexity of cardioversion will improve the efficiency of the procedure for patients and healthcare systems.

**Funding:** None to disclose

**Trial Registration:** ACTRN12620001028998

## INTRODUCTION

### Atrial Fibrillation and Rhythm Control

Atrial fibrillation (AF) is the most common sustained arrythmia worldwide. It will soon represent a public health epidemic and a growing cause of morbidity in millions of individuals. (1, 2) Atrial fibrillation is associated with worse quality of life which can improve after medical therapy. (3) There appear to be significant reductions in mortality with rhythm control for those with AF and heart failure. (4, 5) The East-AF Net-4 trial demonstrated a reduced composite endpoint of cardiovascular death, stroke, heart failure hospitalisation, and acute coronary syndrome with early rhythm control for AF. (6), Therefore, improvements to rhythm control strategies are important to individual patients and to the wider healthcare community.

### Direct Current Cardioversion for Atrial Fibrillation

Direct current cardioversion (DCCV) is an established method of restoring sinus rhythm in those with AF. The success of cardioversion is reported in the literature to be between 70-90%. (7) Biphasic defibrillation improves the success of cardioversion with higher first shock efficacy and decreased energy requirements. (8) Despite this, a significant number of patients fail cardioversion. The main predictors of cardioversion failure are increased body mass index, male sex and higher transthoracic impedance. (9, 10)

### Chest Pressure during Cardioversion

Chest pressure at the time of shock delivery has been shown to decrease transthoracic impedance and increase cardiac energy delivery. (11, 12) Therefore, chest pressure during DCCV could be a promising method to increase the efficacy of electrical cardioversion. While many take care to avoid patient contact during cardioversion, the practical risk of receiving a shock if precautions are taken appears low. (13) A single center randomised trial of 100 patients suggested benefit of chest pressure for reversion to sinus rhythm with improved cardioversion success. (14) This trial started with a low dose of 50 joules, below the guideline recommended starting energy levels of 150 joules. (15)

### Trial Aims and Hypothesis

We aimed to assess the benefit of manual chest pressure for direct current cardioversion for atrial fibrillation. We hypothesised that the routine application of chest pressure would be associated with lower total energy requirements, without increased risk of complications to the patient or proceduralist.

## METHODOLOGY

### Trial Design

This was a multicenter, randomised, investigator-initiated, patient and analysis-blinded trial assessing the utility of chest pressure during direct current cardioversion for atrial fibrillation. The methodology of this trial has been published previously. (16) Ethics approval was obtained from the Hunter New England Research Ethics Committee. This trial was prospectively registered on Australia New Zealand Clinical Trials Registry (ACTRN12620001028998). Inclusion criteria were confirmed AF on electrocardiogram, age greater than 18 years of age, a minimum of three weeks of therapeutic anticoagulation or transoesophageal echocardiography excluding left atrial thrombus, and the ability to provide informed consent. Exclusion criteria were other atrial arrhythmias (atrial flutter or atrial tachycardia), those who were pregnant or breast feeding and those where anti-coagulation was contraindicated. Given the intervention, proceduralists were aware of group assignments. Randomisation occurred via an online tool (Sealed Envelope - https://www.sealedenvelope.com). Patients and the public were not involved in the design, conduct or reporting of the research.

### Intervention and Setting

Patients in control and intervention arms could receive up to four sequential shocks: 150J, 200J, 360J, and 360J of biphasic energy. The control arm received chest pressure with the final shock. The intervention arm received chest pressure from the first shock (see Figure 1). Patients were assigned to control and intervention in a 1:1 fashion. Recruitment occurred across three sites in New South Wales, Australia.

**Figure 1.**
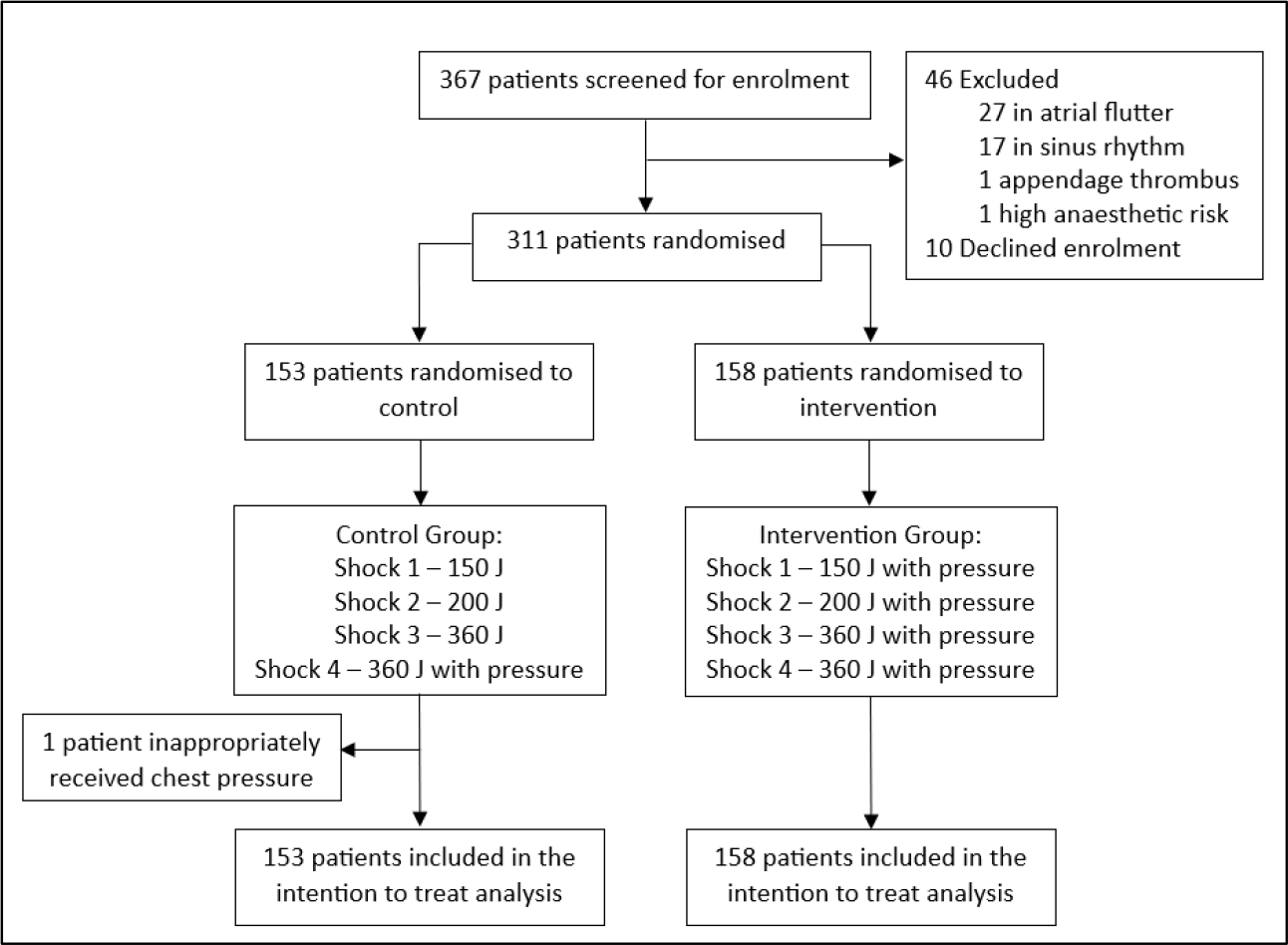
Consort diagram of screening and randomisation.

### Procedural Approach

Written informed consent was gained from all participants. The recommended anaesthetic regimen was weight-based dosing of propofol ± midazolam titrated to effect. Defibrillator pads were placed in an anterior-posterior position. Manual pressure was provided by the proceduralist wearing plastic gloves with a folded towel on the anterior chest with the patient supine (see Figure 2). In our experience, providing chest pressure is a dynamic process, depending on patient size and chest compliance. Therefore, stipulating a set pressure for every patient would not be ideal. Pressure was not standardised or measured during the trial. To provide an estimate of pressure, prior to trial initiation, four cardiology trainees performed 36 applications of blinded simulated pressure with an average of 25.5 ± 2.6kg.

**Figure 2.**
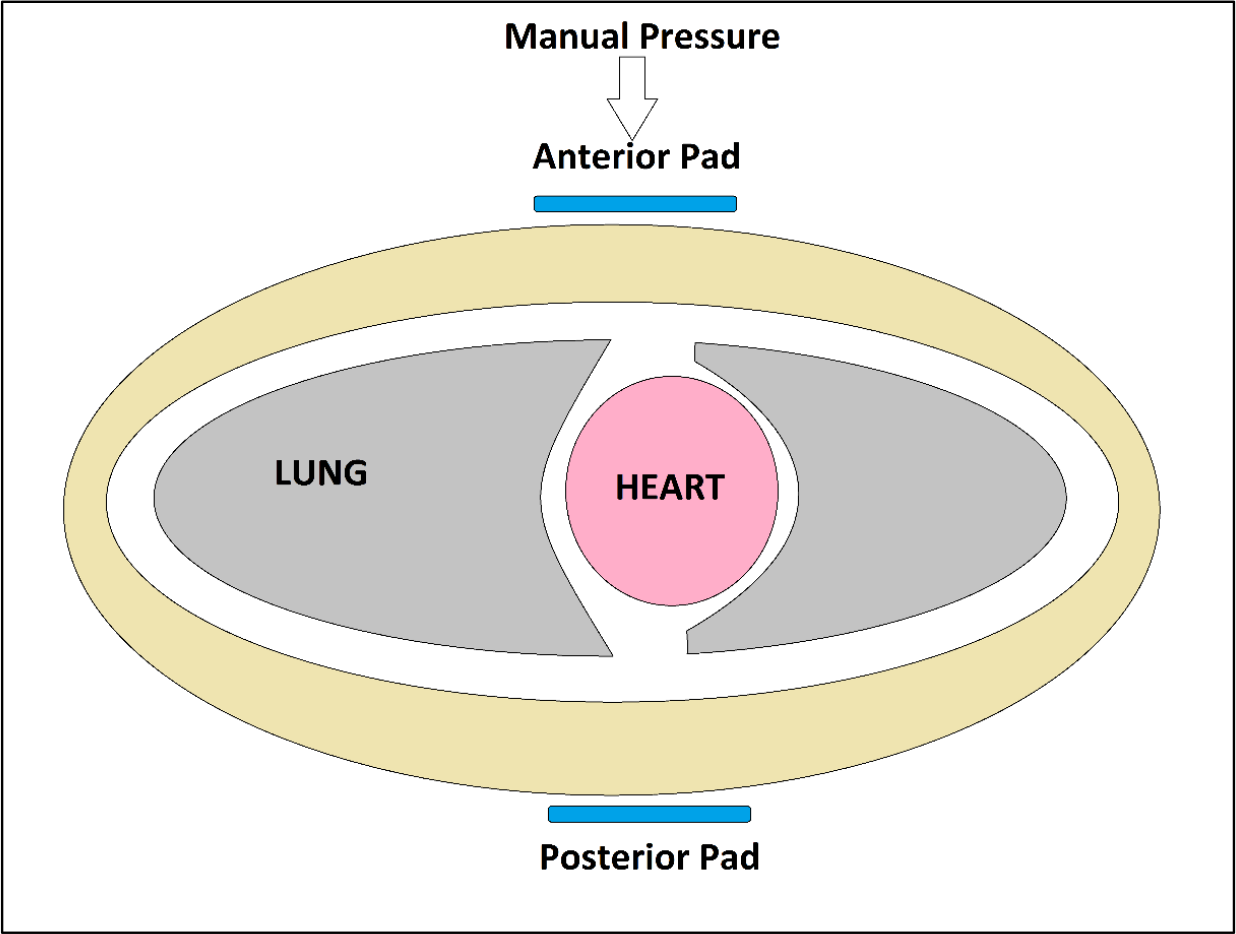
Diagram of chest pressure technique.

### Endpoints

Successful cardioversion was defined as achieving sinus rhythm at one minute post. The primary endpoint was total energy provided per patient encounter. Secondary endpoints included first shock success, transthoracic impedance (provided on the printed telemetry strip), successful cardioversion and sinus rhythm at 30 minutes post procedure. Safety outcomes were patient reported chest pain, and the incidence of shock to proceduralist. Pre-specified post-hoc analyses included stratifying by body mass index, patient age, left atrial volume index and medical therapy (rate and rhythm control).

### Statistical Analysis and Data Sharing

Power Calculation: Based on an audit of cardioversions at our center, the mean energy provided per encounter was 280 ± 188 joules. Assuming a mean reduction of 60 joules in the intervention group (a third of the standard deviation), a sample size of 308 would be required with an alpha of 0.05 and 80% power.

Statistical Analysis: Continuous variables are reported as mean ± standard deviation. Data distribution was determined by the Shapiro-Wilks test. Normally distributed variables were compared using the paired Student’s t-test. Non-parametric data were compared using the Mann-Whitney U test. All tests are two-tailed. A p-value of <0.05 was considered statistically significant. The intention-to-treat analysis was performed blinded to group assignment. To facilitate collaboration, de-identified patient level data is available to researchers upon reasonable request.

## RESULTS

### Recruitment and Baseline Characteristics

Of 367 patients screened, 311 (85%) were randomised with 153 in the control arm and 158 in the intervention arm (see Figure 1). The average age was 66.3 ± 10.9 years with an average body mass index of 31.8 ± 6.6 kg/m^2^. Baseline characteristics between groups were well matched (see Table 1). The median time from diagnosis of atrial fibrillation to cardioversion was six months. Most patients were receiving rhythm control therapy (51%) with 43% on rate control.

**Table 1.** Baseline Characteristics.

| DEMOGRAPHICS | TOTAL | CONTROL | INTERVENTION | P VALUE |
| --- | --- | --- | --- | --- |
| Number | 311 | 153 | 158 |  |
| Age (years) <sup>¥</sup> | 66.3 ± 10.9 | 66.5 ± 11.3 | 66.1 ± 10.5 | 0.33 |
| Female Sex (n [%]) | 77 (24.7%) | 37 (24.2%) | 40 (25.3%) | 0.82 |
| Body Mass Index (kg/m<sup>2</sup>) <sup>¥</sup> | 31.8 ± 6.6 | 31.1 ± 6.1 | 32.6 ± 7.0 | 0.10 |
| Est Glomerular Filtration Rate <sup>¥</sup> | 70.0 ± 18.2 | 70.7 ± 17.5 | 69.3 ± 18.9 | 0.53 |
| AF Diagnosis to DCCV (months) <sup>£</sup> | 6 (3-24) | 6 (3-33) | 6 (3-24) | 0.71 |
| SD of Alcohol <sup>¥</sup> | 7.3 ± 12.6 | 8.4 ± 12.9 | 6.4 ± 12.3 | 0.36 |
| Obstructive Sleep Apnoea (% [n]) | 24.8% (77) | 22.9% (35) | 26.6% (42) | 0.57 |
| COPD (% [n]) | 8.9% (27) | 8.5% (13) | 8.9% (14) | 0.91 |
| Rhythm Control (% [n]) | 50.8% (158) | 51.6% (79) | 50% (79) | 0.77 |
| Rate Control (% [n]) | 42.8% (133) | 39.9% (61) | 45.6% (72) | 0.31 |
| LV Ejection Fraction (% [n]) <sup>¥</sup> | 47.6 ± 13.5 | 48.8 ± 13.1 | 46.6 ± 13.8 | 0.30 |
| Left Atrial Volume Index (% [n]) <sup>¥</sup> | 47.5 ± 14.7 | 46.6 ± 13.9 | 48.3 ± 15.5 | 0.46 |
| Est Estimated, AF atrial fibrillation, DCCV direct current cardioversion, SD Standard drink, COPD chronic obstructive pulmonary disease, LV left ventricular, ¥ Mean with standard deviation, £ Median with interquartile range. |  |  |  |  |

### Efficacy and Safety Outcomes

The primary outcome was available in all participants. There was no difference in total joules applied per encounter in the control arm versus intervention arm, 356.4 ± 301 vs 413.8 ± 347 (P=0.25). There was no difference in first shock success, total shocks provided, average impedance and successful cardioversion (See Table 2).

**Table 2.** Efficacy and Safety Endpoints.

| EFFICACY ENDPOINTS | CONTROL | INTERVENTION | P VALUE |
| --- | --- | --- | --- |
| Total Energy (Joules) <sup>¥</sup> | 356.4 ± 301 | 413.8 ± 347 | 0.25 |
| Total Shocks (n) <sup>¥</sup> | 1.8 ± 1.0 | 1.8 ± 1.1 | 0.21 |
| Average Impedance (ohms) <sup>¥</sup> | 81.8 ± 19.1 | 78.7 ± 17.7 | 0.20 |
| First Shock Success (% [n]) | 59.5% (91) | 52.5% (83) | 0.22 |
| Cardioversion Success (% [n]) | 94.1% (144) | 91.5% (140) | 0.08 |
| Sinus at 30 Minutes (% [n]) | 92.1% (141) | 86.7% (137) | 0.12 |
| **SAFETY ENDPOINTS** |  |  |  |
| Chest Pain Post (% [n]) | 1.3% (2) | 1.9% (3) | NS |
| Pad Related Burn (% [n]) | 0.7% (1) | 0% (0) | NS |
| Shock to Proceduralist (% [n]) | 0% (0) | 0% (0) | NS |
| NS non-significant. ¥ Mean with standard deviation |  |  |  |

Adverse events were low and did not differ between groups (see Table 2). There were no shocks experienced by any proceduralist. Transient bradycardia and hypotension occurred in two patients in control and one patient in the intervention arm. There was one aspiration event during recovery (requiring intensive care unit admission and non-invasive ventilation) in an individual within the intervention arm. There were no ischaemic cerebrovascular events noted with follow-up until discharge.

### Subgroup Analysis

Pre-specified analyses were performed based on body mass index, left atrial volume index, age and baseline medical therapy (rate control and rhythm control). Response to chest pressure during cardioversion was not influenced by these factors (see Table 3). Moreover, in a post-hoc analysis assessing only the first three shocks (excluding the fourth/final shock) there was no difference in total joules between control and intervention arms (323 ± 232 verse 354 ± 245 respectively, P=0.26) and improved overall success within the control group (91.5% verse 84.2%, P=0.049). In all these pre-specified analyses the effect size favoured control rather than intervention, though in the context of multiple comparisons, no inference can be drawn.

**Table 3.** Pre-specified Subgroup Analysis of the Primary Outcome, Total Joules.

|  | CONTROL | INTERVENTION | P VALUE |
| --- | --- | --- | --- |
| **BODY MASS INDEX** |  |  |  |
| Body Mass Index ($\geq 30 \text{ kg/m}^2$)<sup>¥</sup> | 378 ± 311 | 457 ± 356 | 0.19 |
| Body Mass Index ($\geq 35 \text{ kg/m}^2$)<sup>¥</sup> | 466 ± 358 | 539 ± 391 | 0.52 |
| Body Mass Index ($\geq 40 \text{ kg/m}^2$)<sup>¥</sup> | 558 ± 382 | 545 ± 387 | 0.85 |
| **LEFT ATRIAL VOLUME INDEX** |  |  |  |
| Left Atrial Volume Index ($\geq 30 \text{ mL/m}^2$)<sup>¥</sup> | 345 ± 297 | 435 ± 357 | 0.19 |
| Left Atrial Volume Index ($\geq 40 \text{ mL/m}^2$)<sup>¥</sup> | 342 ± 284 | 421 ± 348 | 1.0 |
| Left Atrial Volume Index ($\geq 50 \text{ mL/m}^2$)<sup>¥</sup> | 257 ± 185 | 402 ± 358 | 0.21 |
| **AGE** |  |  |  |
| Age ($\geq 50$ years)<sup>¥</sup> | 352 ± 297 | 414 ± 351 | 0.27 |
| Age ($\geq 60$ years)<sup>¥</sup> | 356 ± 300 | 418 ± 347 | 0.22 |
| Age ($\geq 70$ years)<sup>¥</sup> | 314 ± 264 | 395 ± 352 | 0.35 |
| **MEDICAL THERAPY** |  |  |  |
| Rate Control Therapy | 372 ± 318 | 421 ± 347 | 0.34 |
| Rhythm Control Therapy | 332 ± 272 | 422 ± 353 | 0.23 |
| ¥ Mean with standard deviation |  |  |  |

## DISCUSSION

In this randomised control trial, the routine application of chest pressure did not reduce energy requirements for cardioversion. Chest pressure also did not improve first shock success or reduce transthoracic impedance during cardioversion. Safety endpoints and major adverse events were similarly low between the groups. Given both groups could receive a 360J shock with chest pressure, overall procedural success was not expected to be different.

The lack of efficacy is notable, and these results were unexpected based on prior studies. This may be due to several reasons. First, the higher starting energies used may be at a threshold where chest pressure provided no added benefit. Secondly, data for chest pressure reducing transthoracic impedance is mostly sourced from manual paddles. (11, 12, 17) There is evidence to suggest that the majority of transthoracic impedance is from the paddle-skin interface. (18, 19) In the modern era, less transthoracic impedance is observed with the use of adhesive pads. Thirdly, while the method of pressure application was standardised, it was not measured and may have varied between operators. Insufficient pressure could explain the lack of difference in impedance and efficacy between groups, but we believe this is unlikely. The optimal pressure provision for impedance reduction is reported as 8 kilograms on each paddle, and a total of 16 kilograms across the chest. (17, 19) Prior to undertaking our trial, 36 blinded applications of chest pressure were performed by 4 operators. The average pressure provided was 25.5 ± 2.6kg, greater than that recommended by the data. The narrow standard deviation suggests that operators provided similar pressure. Moreover, we suggest our trial is externally valid and generalizable, demonstrating that in a real-world setting the routine delivery of chest pressure did not improve cardioversion efficacy.

We found that first shock success with 150 joules was low, at only 56%. Those with greater body mass index were more likely to require greater total energies. Though not directly tested in this trial, those with higher BMI’s may gain greater benefit from higher starting energies.

Though the average energy provided per encounter was higher than used in the sample size calculation, this trial still had adequate power. The mean energy provided within the control group was 356J ± 301J. Assuming a reduction of one third the standard deviation in the intervention arm (100 Joules) 284 total patients would have been required to detect a difference with a two-sided alpha of 0.05 and a power of 80%. This is a smaller sample size than the number enrolled in this trial.

There were no episodes of shock experienced by the proceduralists. If precautions are taken similar to our practice (plastic gloves and a towel), the likelihood of a shock experienced by an individual performing chest pressure/compressions seems low. This is contrary to the traditional teaching that all contact must be avoided with the patient when cardioversion occurs. This may have implications during advanced life support for shockable rhythms.

There are limitations to this trial. While the method of applying chest pressure was standardised for the trial, the amount of pressure was not measured and may vary according to operator. Though pre-specified subgroup analyses did not demonstrate any baseline characteristic interaction with treatment effect, our study lacked power to determine whether subgroups may benefit from the intervention.

## CONCLUSION

The routine application of chest pressure did not result in reduced energy requirements for cardioversion of atrial fibrillation in this randomised multi-center trial. The application of chest pressure during cardioversion was safe and feasible though not associated with benefit when administered routinely.

## Data Availability

De-identified patient level data will be available upon reasonable request after review of analysis plan.

## Notes

**FUNDING** AL Sverdlov is supported by the National Heart Foundation of Australia Future Leader Fellowship (award ID 106025) and the NSW Health Cardiovascular Capacity Building Grant.

**CONFLICTS OF INTEREST** None declared.

### Competing Interest Statement

The authors have declared no competing interest.

### Clinical Trial

Australia New Zealand Clinical Trials Registry ACTRN12620001028998

### Clinical Protocols

https://openheart.bmj.com/content/8/2/e001739

### Author Declarations

Hunter Research Ethics Committee

